# Circumferential aneurysm wall enhancement predicts recanalization after stent-assisted coiling in small unruptured intracranial aneurysms

**DOI:** 10.1101/2025.06.24.25330199

**Authors:** Qiu-Yi Jiang, Xin-Lei Ruan, Rui Chen, Zhong-Song Shi

**Affiliations:** Department of Neurosurgery (Q-YJ, X-LR, RC, Z-SS), Sun Yat-sen Memorial Hospital, Sun Yat-sen University, Guangzhou, China; RNA Biomedical Institute (Z-SS), Sun Yat-sen Memorial Hospital, Sun Yat-sen University, Guangzhou, China; Guangdong Province Key Laboratory of Brain Function and Disease (Z-SS), Sun Yat-sen University, Guangzhou, China

**Keywords:** Unruptured intracranial aneurysm, Endovascular treatment, Recanalization, Vessel wall imaging, Aneurysm wall enhancement

## Abstract

**Background:** Circumferential aneurysm wall enhancement (CAWE) on high-resolution vessel wall imaging (HR-VWI) as a vessel wall inflammation marker is associated with the instability of unruptured intracranial aneurysms (UIAs) and recanalization after endovascular treatment. This study evaluates the association of CAWE with recanalization of small UIAs (<10 mm) treated with stent-assisted coiling (SAC) or coiling alone and aims to develop a prediction model for recanalization based on CAWE.

**Methods:** We analyzed patients with saccular UIAs who underwent 3T HR-VWI and were treated with SAC or coiling alone between October 2018 and May 2024. A 4-grade scale assessed aneurysm wall enhancement (none, focal thick wall enhancement, thin CAWE, thick CAWE). The aneurysm-to-pituitary stalk contrast ratio (CRstalk) quantified enhancement. We investigated the relationship between CAWE and recanalization and developed a recanalization prediction model.

**Results:** Sixty-five patients with 69 small saccular UIAs were included; 11 aneurysms (15.9%) exhibited CAWE, and 10 aneurysms (14.5%) had a CRstalk ≥ 0.5. Sixty aneurysms received SAC. Recanalization occurred in 14 of 69 aneurysms (20.3%), assessed by digital subtraction angiography follow-up at 12.6 months. Multivariate analysis revealed that smoking, aneurysm size, CAWE, and CRstalk ≥ 0.5 predicted recanalization. A scoring prediction model was created using aneurysm size, treatment, embolization occlusion, and CAWE, with scores ranging from 0 to 6, where scores ≥ 3 indicated high risk and a C-statistic of 0.892 demonstrated excellent discrimination.

**Conclusions:** CAWE on HR-VWI is a significant imaging marker for predicting recanalization in small UIAs undergoing SAC. The proposed recanalization risk scale needs further validation in larger studies.

## Introduction

The rupture of intracranial aneurysms leading to subarachnoid hemorrhage can be effectively addressed through endovascular treatment or surgical clipping. The International Subarachnoid Aneurysm Trial study indicates that over 80 percent of ruptured aneurysms are small-sized (less than 10 mm).^1^ Unruptured intracranial aneurysms (UIAs) of small size demonstrate a low risk of rupture and growth.^2–5^ Notably, tiny UIAs (3 mm or smaller) are generally not recommended for preventative treatment.^6^ In the last twenty years, more intracranial aneurysms have been treated using safer endovascular techniques, and the size of treated UIAs has decreased over time.^7, 8^ The modalities, including endovascular coiling, stent-assisted coiling (SAC), balloon-assisted coiling, flow diverters, and Woven EndoBridge devices, are effective treatment options for small and medium-sized UIAs with wide necks.^9–13^ The use of flow diverters for small UIAs carries a risk of 2% for major stroke or neurological death.^10–12^ Coiling alone and coiling with stent or balloon assistance have limitations due to the associated rates of aneurysm recanalization.^9, 14, 15^ Several factors related to patients and aneurysm morphology are linked to recanalization after coiling, such as smoking, aneurysm size, neck size, and aneurysm location. SAC lowers recanalization rates without increasing complication risks.^14–16^

High-resolution vessel wall imaging (HR-VWI) has become an essential MRI technique for evaluating the thickness and enhancement of aneurysms.^17^ Aneurysm wall enhancement (AWE) is a crucial imaging biomarker that indicates the histological features of degenerative changes, neovascularization, and inflammation within the aneurysm wall.^18^ Circumferential AWE (CAWE) and quantitative analysis with a higher contrast ratio measurement are more strongly associated with aneurysm instability, an important indicator for preventative treatments in UIAs to help reduce their significant risk of rupture.^19–22^ Preoperative AWE in saccular UIAs predicts clinical outcome at 6-month follow-up after surgical clipping.^23^ CAWE before endovascular treatment can predict aneurysm recanalization or enlargement of unruptured vertebrobasilar dissection aneurysms following SAC or stenting alone.^24^ In the case of saccular UIAs, CAWE prior to endovascular coiling is also associated with aneurysm recanalization.^25^ There is limited evidence regarding the prediction of pretreated AWE in recanalization following SAC in small UIAs (<10 mm). This study aimed to investigate the association of CAWE and quantitatively measured enhancement in 3T HR-VWI with recanalization of small UIAs treated with SAC and to develop a prediction model for recanalization based on CAWE.

## Methods

### Study population

We selected consecutive patients with UIA for initial preventative treatment using endovascular and surgical techniques from the HR-VWI aneurysm database at our institution between October 2018 and May 2024. The study included adult patients (older than 18 years) with small saccular UIAs (ranging from 3 to 10 mm) treated with endovascular coiling alone or with SAC. We excluded patients with aneurysms in the extracranial or cavernous sinus sections of the internal carotid artery. All patients with aneurysms had an identified MR angiography and a favorable quality of HR-VWI images for assessment. Furthermore, all participants received digital subtraction angiography follow-up at least three months after endovascular treatment. The study received approval from the Ethics Committee at our institution.

We collected the data on demographics, laboratory examination, radiological, and clinical characteristics. The morphological factors of aneurysm included the maximum aneurysm size, neck size, location, size ratio, aspect ratio, dome-to-neck ratio, and bifurcation or sidewall pattern. We assessed the risk of aneurysm rupture and growth using the PHASES and ELAPSS scores.

### Aneurysm wall enhancement on HR-VWI

Patients underwent brain MR for HR-VWI on a 3.0T MRI scanner (Achieva TX, Philips Healthcare, Best, the Netherlands) with a 32-channel head coil. The HR-VWI protocol with precontrast and postcontrast sequences was described in our previous study. The assessment of AWE in the pre-treatment MR was conducted using both a subjective AWE scale and an objective measurement of the signal intensity of the aneurysm wall, as described in earlier studies.^19, 20, 21, 26^ A four-grade scale was used to evaluate AWE. Grade 0: No definite or questionable enhancement compared with the pre-contrast sequence. Grade 1: Focal thick enhancement with thickness > 1 mm. Grade 2: Thin, circumferential enhancement of the entire wall with thickness ≤ 1 mm. Grade 3: Thick, circumferential enhancement of the entire wall with thickness > 1 mm, or thin circumferential enhancement of the entire wall with focal enhancement thickness > 1 mm. AWE grades 2 and 3 were classified as CAWE.^19^ For quantitative analysis, the maximum signal intensity of the aneurysmal wall (SIwall) and the pituitary stalk (SIstalk) was measured. The aneurysm-to-pituitary stalk contrast ratio (CRstalk) was then calculated to reflect the degree of AWE, using the formula: CRstalk = SIwall/SIstalk.^20, 21, 26^

### Endovascular treatment and angiographic follow-up

Patients underwent endovascular treatment using coiling, with or without stent placement, while under general anesthesia. SAC was chosen when an aneurysm had a wide neck (≥4 mm) or a dome-to-neck ratio <2. Bare platinum coils and Enterprise-2 stents (Codman Neuro) were utilized in this study. Patients who received SAC were prescribed dual antiplatelet regimens, including Aspirin, clopidogrel, and platelet inhibition testing.

The modified Raymond-Roy classification (MRRC) was utilized to evaluate the degree of aneurysm embolization, assigning grades I, II, and III (with subcategories IIIa and IIIb).^27^ MRRC grade I indicates complete occlusion, while grades I and II are considered satisfactory occlusion based on the immediate post-embolization angiogram. The modified Rankin Scale was assessed at discharge, outpatient visits, and inpatient clinics. Follow-up DSA was conducted at 3-6 months, 12 months, and 24 months post-treatment. Evaluations focused on coil compaction in the sac and neck and aneurysm regrowth. An increase in the MRRC grade on follow-up angiography compared to immediate post-treatment angiograms is considered an aneurysm recanalization.^14, 15^

## Data analysis

R version 4.2.2 was used for statistical analysis. Two researchers, blinded to the clinical and radiological data, independently assessed the AWE and MRRC pattern. The inter-rater agreement was measured using the Cohen κ coefficient and intra-class correlation coefficient. A third researcher resolved the disagreement. Collinearity among variables was evaluated using the variance inflation factor (VIF > 2.5) to mitigate multicollinearity. We analyzed the continuous variables using Student’s t-test or Mann-Whitney U test, and categorical variables by chi-square or Fisher’s exact test. Then, we performed the univariate and multivariate logistic regression analyses to determine the independent factors related to aneurysm recanalization. A p-value < 0.05 was considered statistically significant.

By integrating the findings from this study with previous research and clinical practice experience, we identified predictive factors to construct a multivariable logistic regression model. Following the established method, we converted aneurysm size into a categorical variable to create a clinically applicable scoring system, assigning a value of 1 to the minimum non-zero regression coefficient. Next, we calculated the ratio of each predictor’s regression coefficient to this baseline value, rounded it to the nearest integer, and assigned values accordingly to develop an aneurysm recanalization risk score. We meticulously evaluated the performance of this model using receiver operating characteristic (ROC) curve analysis, a confusion matrix, and a calibration curve. The critical threshold for risk stratification was determined based on the optimal cut-off value identified from the ROC curve. We further elucidated the multivariable logistic regression model using the SHapley Additive exPlanations (SHAP) method to enhance the interpretability of the model.^28^ **Results**

### Patient, aneurysm, and HR-VWI characteristics

Two hundred eight consecutive patients with UIAs receiving pre-treatment HR-VWI assessment and endovascular treatment were identified during the study period. Sixty-five patients with 69 saccular UIAs were included in this study according to the predefined eligibility criteria. The mean age was 57.5 ± 10.9 years, and 39 (58%) were female. The mean size of aneurysms was 5.7 ± 1.8 mm, with 26 (36%) presenting with irregular shapes. All aneurysms had a maximum size of less than 10 mm, with 3.0-4.9 mm size in 26 aneurysms, 5.0-6.9 mm in 29, and 7.0-9.9 mm in 14. Twenty-five aneurysms had a wide-neck (≥4.0 mm), and 65 aneurysms had a dome-to-neck ratio <2. Forty-seven (66.2%) aneurysms were located in the internal carotid artery, 12 (16.9%) aneurysms were in the middle cerebral artery, 5 (7.0%) aneurysms were in the anterior communicating artery, 4 (5.6%) in the posterior communicating artery, and 3 (4.2%) in the posterior circulation. The PHASES score ranged from 0 to 8, and the ELAPSS score ranged from 5 to 23.

A 4-grade scale AWE at 3T HR-VWI showed 47 as grade 0, 11 as grade 1, 4 as grade 2, and 7 as grade 3. The pattern of CAWE included AWE grades 2 and 3, which were observed in 11 aneurysms (15.9%). The interreader agreement for the identification of AWE was excellent, with κ=0.90. The mean value of CRstalk was 0.33 ± 0.16 in this study. The CRstalk value ≥ 0.5 related to aneurysm instability in saccular UIAs was previously shown. Ten of 69 aneurysms (14.5%) had a CRstalk ≥ 0.5. The interreader agreement for the measurement of CRstalk was excellent, with intra-class correlation coefficient=0.99. The characteristics of the patient, aneurysm, and image are shown in Table 1.

**Table 1.** Characteristics of unruptured intracranial aneurysms.

|  | **Total (n=69)** | **Recanalization**<br>**(n=14)** | **No**<br>**Recanalization**<br>**(n=55)** | **P Value** |
| --- | --- | --- | --- | --- |
| Age (yr) | 57.3 ± 10.8 | 57.1 ± 12.5 | 57.4 ± 10.5 | 0.946 |
| Female | 40 (58.0%) | 4 (28.6%) | 36 (65.5%) | 0.017 |
| Male | 29 (42.0%) | 10 (71.4%) | 19 (34.5%) | 0.017 |
| Hypertension | 32 (46.4%) | 6 (42.9%) | 26 (47.3%) | 1.000 |
| Diabetes | 10 (14.5%) | 2 (14.3%) | 8 (14.5%) | 1.000 |
| Dyslipidemia | 5 (7.2%) | 2 (14.3%) | 3 (5.5%) | 0.266 |
| Smoking | 13 (18.8%) | 6 (42.9%) | 7 (12.7%) | 0.028 |
| Aneurysm size (mm) | 5.7 ± 1.8 | 7.5 ± 2.0 | 5.2 ± 1.5 | <0.001 |
| 3.0-4.9 mm | 26 (37.7%) | 2 (14.3%) | 24 (43.6%) |  |
| 5.0-6.9 mm | 29 (42.0%) | 3 (21.4%) | 26 (47.3%) |  |
| 7.0-9.9 mm | 14 (20.3%) | 9 (64.3%) | 5 (9.1%) |  |
| Neck size ≥4.0 mm | 25 (36.2%) | 8 (57.1%) | 17 (30.9%) | 0.131 |
| Site of aneurysm |  |  |  | 0.022 |
| ICA | 46 (66.7%) | 5 (35.7%) | 41 (74.5%) |  |
| MCA | 11 (15.9%) | 4 (28.6%) | 7 (12.7%) |  |
| ACoM | 5 (7.2%) | 2 (14.3%) | 3 (5.5%) |  |
| PCoM | 4 (5.8%) | 1 (7.1%) | 3 (5.5%) |  |
| PC | 3 (4.3%) | 2 (14.3%) | 1 (1.8%) |  |
| Bifurcation aneurysm | 22 (31.9%) | 8 (57.1%) | 14 (25.5%) | 0.051 |
| PHASES score | 1.0 (0.0-4.0) | 5.0 (3.0-6.0) | 1.0 (0.0-3.0) | <0.001 |
| ELAPSS score | 12.0 (9.0-15.0) | 15.5 (14.0-19.0) | 11.0 (7.5-15.0) | 0.001 |
| AWE |  |  |  | <0.001 |
| Grade 0 | 47 (68.1%) | 3 (21.4%) | 44 (80.0%) |  |
| Grade 1 | 11 (15.9%) | 3 (21.4%) | 8 (14.5%) |  |
| Grade 2 | 4 (5.8%) | 2 (14.3%) | 2 (3.6%) |  |
| Grade 3 | 7 (10.1%) | 6 (42.9%) | 1 (1.8%) |  |
| CAWE, grade 2 and 3 | 11 (15.9%) | 8 (57.1%) | 3 (5.5%) | <0.001 |
| CR stalk | 0.33 ± 0.16 | 0.50 ± 0.16 | 0.28 ± 0.12 | <0.001 |
| Endovascular treatment |  |  |  |  |
| Stent-assisted coiling | 60 (87.0%) | 9 (64.3%) | 51 (92.7%) | 0.014 |
| Coiling alone | 9 (13.0%) | 5 (35.7%) | 4 (7.3%) | 0.014 |
| Embolization occlusion |  |  |  | 0.032 |
| MRRC grade I | 48 (69.6%) | 7 (50.0%) | 41 (74.5%) |  |
| MRRC grade II | 16 (23.2%) | 7 (50.0%) | 9 (16.4%) |  |
| MRRC grade IIIa+IIIb | 5 (7.2%) | 0 (0.0%) | 5 (9.1%) |  |
| Coil compaction | 13 (18.8%) | 11 (78.6%) | 2 (3.6%) | <0.001 |

ACoM, indicates anterior communicating artery; AWE, aneurysm wall enhancement; CAWE, circumferential aneurysm wall enhancement; CR, contrast ratio; ICA, internal carotid artery; MCA, middle cerebral artery; MRRC, Modified Raymond-Roy Classification; PComA, posterior communicating artery; PC, posterior circulation.

### Aneurysm recanalization after endovascular treatment

Sixty aneurysms received SAC, and nine aneurysms were treated by coiling only. Complete occlusion was shown as MRRC grade I in 48 aneurysms at the immediate embolization angiogram. The satisfactory embolization occlusion was achieved in 54 aneurysms, with 48 in MRRC grade I and 16 in MRRC grade II. There were no ischemic or hemorrhagic complications related to endovascular treatment. Aneurysm recanalization was detected in 14 of 69 (20.3%), as shown by digital subtraction angiography follow-up at 12.6 (3-23) months. Of these 14 recanalized aneurysms, 13 exhibited coil compaction (Figure 1). All 65 patients had a favorable clinical outcome with an mRS scale of less than 1 at 90 days and are monitored regularly.

### CAWE and CRstalk as predictors of aneurysm recanalization

In the univariable analysis showed in Table 1, recanalized aneurysms were more prevalent in male patients (71.4% vs. 34.5%, p=0.017) and smokers (42.9% vs. 12.7%, p=0.028). Recanalized aneurysms were significantly larger (7.5 mm vs. 5.2 mm, p<0.001) and were less frequently located in the internal carotid artery (35.7% vs. 74.5%, p=0.007). Higher PHASES and ELAPSS scores were observed in the recanalized aneurysms. The AWE pattern was more pronounced in recanalized aneurysms compared to non-recanalized ones (all AWE, 78.6% vs. 18.2%, p<0.001; all CAWE, 57.1% vs. 5.5%, p<0.001; CAWE grade 3, 42.9% vs. 1.8%, p<0.001). A higher mean CRstalk value (0.50 vs. 0.28, p<0.001) and higher percent of CRstalk ≥ 0.5 (50.0% vs. 5.5%, p<0.001) were exhibited in the recanalized aneurysms. Recanalized aneurysms were more frequently treated with coiling alone (35.7% vs. 7.3%, p=0.014). Coil compaction occurred significantly more often in recanalized aneurysms (78.6% vs. 3.6%, p<0.001).

In the multivariate analysis, after adjusting variables of smoking, aneurysm size, location in the internal carotid artery, complete occlusion at immediate embolization, coiling alone, CAWE with grade 2 and grade 3, and CRstalk ≥ 0.5, the results showed that smoking (OR 10.26, 95% CI 1.51-96.79, p=0.022), aneurysm size (OR 2.19, 95% CI 1.28-4.53, p=0.012), CAWE with grade 2 and grade 3 (OR 28.32, 95% CI 3.87-364.92, p=0.003), CAWE with grade 3 (OR 85.63, 95% CI 6.23-3313.51, p=0.004), CRstalk ≥ 0.5 (OR 11.09, 95% CI 1.51-116.15, p=0.024) were independently associated with recanalization after endovascular treatment (Table 2). In the univariable analysis of 60 aneurysms treated with SAC, the above factors were also related to aneurysm recanalization. Aneurysm size, CAWE with grades 2 and 3, CAWE with grade 3, and CRstalk ≥ 0.5 remained independent risk factors for recanalization (Table 3).

**Table 2.** Multivariable logistic regression analysis of risk factors associated with aneurysm recanalization after stent-assisted coiling and coiling alone.

| Variable | Model 1 |  | Model 2 |  | Model 3 |  |
| --- | --- | --- | --- | --- | --- | --- |
|  | Odds ratio<br>(95% CI) | *P* value | Odds ratio<br>(95% CI) | *P* value | Odds ratio<br>(95% CI) | *P* value |
| Smoking |  |  | 10.26<br>(1.51-96.79) | 0.022 | - | - |
| Aneurysm size | 2.19 (1.28-4.53) | 0.012 | 2.16 (1.25-4.44) | 0.014 | 2.19 (1.27-4.53) | 0.013 |
| CAWE, grade 2 and 3 | 28.32<br>(3.87-364.92) | 0.003 | - | - | - | - |
| CAWE, grade 3 | - | - | 85.63<br>(6.23-3313.51) | 0.004 | - | - |
| CR stalk $\geq 0.5$ | - | - | - | - | 11.09<br>(1.51-116.15) | 0.024 |

Abbreviation: CAWE, circumferential aneurysm wall enhancement; CI, confidence interval. CR, contrast ratio.
Model 1 was adjusted for smoking, aneurysm size, aneurysm location in the internal carotid artery, aneurysm complete occlusion at immediate embolization, coiling alone, CAWE with grade 2 and grade 3. Model 2 was adjusted for smoking, aneurysm size, internal carotid artery, complete occlusion, coiling alone, and CAWE with grade 3. Model 3 was adjusted for smoking, aneurysm size, internal carotid artery, complete occlusion, coiling alone, and CR stalk $\geq 0.5$ .

**Table 3.** Multivariable logistic regression analysis of risk factors associated with aneurysm recanalization after the subgroup of stent-assisted coiling.

| Variable | Model 1 |  | Model 2 |  | Model 3 |  |
| --- | --- | --- | --- | --- | --- | --- |
|  | Odds ratio<br>(95% CI) | P value | Odds ratio<br>(95% CI) | P value | Odds ratio<br>(95% CI) | P value |
| Aneurysm size | 2.11 (1.18-4.77) | 0.027 | 2.05 (1.23-4.00) | 0.013 | 1.85 (1.12-3.39) | 0.025 |
| CAWE, grade 2 and 3 | 55.48 (4.72-1425.64) | 0.004 | - | - | - | - |
| CAWE, grade 3 | - | - | 31.75 (1.75-1334.83) | 0.032 | - | - |
| CR stalk $\geq 0.5$ | - | - | - | - | 14.24 (1.25-244.63) | 0.040 |

Abbreviation: CAWE, circumferential aneurysm wall enhancement; CI, confidence interval. CR, contrast ratio.
Model 1 was adjusted for smoking, aneurysm size, aneurysm location in the internal carotid artery, aneurysm complete occlusion at immediate embolization, CAWE with grade 2 and grade 3. Model 2 was adjusted for smoking, aneurysm size, internal carotid artery, complete occlusion, and CAWE with grade 3. Model 3 was adjusted for smoking, aneurysm size, internal carotid artery, complete occlusion, and CR stalk $\geq 0.5$ .

### Recanalization risk score model

Based on the multivariable analyses, a recanalization risk scoring system integrating aneurysm size, CAWE, embolization occlusion, and treatment modality was developed for small UIAs less than 10 mm following SAC and coiling based on the multivariable analyses (Table 4). The total score was 0 to 6 points, with scores ≥ 3 indicating a high risk of recanalization. The recanalization rates at each score level were as follows: 0, 6.3%; 1, 0.0%; 2, 20.0%; 3, 66.7%; 4, 60.0%; 5, 100%. The median risk score was 1.0, and the mean score was 1.60 ± 1.46.

**Table 4.** Aneurysm recanalization risk scale.

| **Factors** | **Points assigned** |
| --- | --- |
| Aneurysm size (mm) |  |
| 3.0-4.9 | 0 |
| 5.0-6.9 | 1 |
| 7.0-10.0 | 2 |
| Aneurysm wall enhancement |  |
| Grade 0-1 | 0 |
| Grade 2-3 | 2 |
| Embolization occlusion |  |
| MRRC I | 0 |
| MRRC II, IIIa, IIIb | 1 |
| Treatment modality |  |
| Stent-assisted coiling | 0 |
| Coiling alone | 1 |

The risk scoring system demonstrated good discriminative ability and adequate calibration, indicating reasonable predictive performance. An area under the curve from ROC analysis was 0.893, with an optimal probability threshold of 0.394 determined by the Youden index (Figure 2A). The model of the confusion matrix achieved a sensitivity of 71.4%, specificity of 94.5%, accuracy of 89.9%, precision of 76.9%, and an F1 score of 74.0% (Figure 2B). Calibration analysis indicated strong concordance between predicted and observed outcomes with the p-value of Hosmer-Lemeshow equal to 0.400 and the Brier score of 0.083 (Figure 2C). Feature importance and the direction of feature effects were visualized using a SHAP beeswarm plot. Ranking of features by their mean absolute SHAP values confirmed aneurysm size as the most influential predictor of recanalization, followed by CAWE, embolization occlusion, and treatment modality (Figure 2D).

## Discussion

Our study found several factors associated with recanalization following SAC and coiling alone for small saccular UIAs less than 10 mm in size with wide necks. These factors include sex, smoking, aneurysm size, location, PHASES score, ELAPSS score, AWE pattern, CRstalk, type of endovascular treatment, embolization occlusion grade, and coil compaction. In our multivariate logistic regression analysis, after adjusting variables such as smoking, aneurysm size, location in the internal carotid artery, complete occlusion at immediate embolization, coiling alone, CAWE with grades 2 and 3, and CRstalk ≥ 0.5, we found that smoking, aneurysm size, CAWE, and CRstalk ≥ 0.5 were independently associated with recanalization. Additionally, we developed a scoring-based prediction model to assess the risk of recanalization using aneurysm size, type of endovascular treatment, embolization occlusion grade, and CAWE. The total score of this predictive model ranges from 0 to 6 points, with scores ≥ 3 indicating a high risk of recanalization. The model demonstrated excellent discriminative ability, achieving a C-statistic of 0.892.

Coiling compaction and aneurysm growth were the main imaging findings for recanalization after coiling embolization.^14, 15^ In our study, over 85% of aneurysms were treated by SAC. Coiling compaction detected in the angiographic follow-up was associated with aneurysm recanalization following SAC, even in aneurysms with initial complete embolization. Previous studies of histopathological examinations of recanalized aneurysms showed fresh thrombus, granulation tissue, and scar tissue in specific areas. Inadequate thrombus fibrosis with insufficient neck endothelialization and impaired smooth muscle cells with inflammatory cells infiltrating the aneurysm wall contribute to recanalization.^29^

The imaging biomarker CAWE histologically reflects the entire aneurysm wall with inflammation.^17, 18^ Our study provides new evidence that pretreatment CAWE can predict aneurysm recanalization following SAC in small-sized saccular UIAs less than 10 mm with wide necks. A recent study reported that CAWE and volume embolization ratio were associated with recanalization after endovascular treatment for saccular UIAs, in which two-thirds of UIAs received coiling alone. Aneurysm recanalization was observed in 18 out of 67 aneurysms by follow-up with magnetic resonance angiography. Of these recanalized aneurysms, 12 exhibited coil compaction, while six experienced growth.^25^ These findings support the potential of pretreatment CAWE as a predictive tool for aneurysm recanalization following SAC or coiling alone.

Enhancement of the aneurysm wall, aneurysm cavity, or stented parent artery wall from the follow-up HR-VWI was also found in aneurysms after endovascular treatment with coiling or flow diverters. These observations, which suggest either aneurysm healing or inflammatory reactions post-endovascular coiling, remain debatable.^30–34^ Recent studies have indicated a correlation between post-treatment AWE and aneurysm recanalization, particularly noting the largest recanalization diameter after initial complete occlusion through coiling, with a mean follow-up period of 43 months.^30^ The relationship between posttreatment AWE and recanalization is notably more pronounced in coiled aneurysms with a maximum diameter exceeding 7.5 mm.^31^ The potential of posttreatment AWE as an indicator of aneurysm instability in coiled aneurysms that have achieved complete occlusion necessitates further investigation. HR-VWI with contrast enhancement recently exited its effectiveness as an invasive tool for assessing aneurysm occlusion and in-stent stenosis after flow diverter treatment.^32, 33^ Additionally, at the one-year follow-up, AWE was observed in nearly 80% of complete occluded aneurysms and 100% of non-occluded aneurysms. Stented parent artery wall enhancement was more announced in aneurysms with AWE.^32^ The occurrence of CAWE was noted in 23.8% of aneurysms before treatment, rising to 38% after flow diverter treatment, with an average follow-up period of 8 months. Posttreatment AWE patterns observed at follow-up did not significantly correlate with pretreatment AWE patterns and aneurysm occlusion in cases treated with flow diverters.^34^

Objective quantification of the signal intensity of the aneurysm wall can provide an accurate assessment of AWE and reduce inconsistencies associated with subjective evaluations.^17, 26, 35^ The CRstalk measurement, which utilizes the maximal signal intensity, is a significant indicator of aneurysm instability.^26^ The value of CRstalk in unstable UIAs was higher than that of stable UIAs.^20^ In our previous study involving 75% saccular aneurysms smaller than 7 mm, a CRstalk value ≥ 0.5 was linked to aneurysm instability in UIAs.^21^ Besides using the subjective AWE scale as in previous studies, we discovered that the CRstalk value in recanalized aneurysms is higher than in non-recanalized aneurysms. A CRstalk value of 0.5 or greater has been identified as a predictor of aneurysm recanalization after SAC. Recent advancements in three-dimensional quantification mapping and histogram analysis have significantly improved the accuracy of evaluating aneurysm walls.^35^ A radiomics composite score provides a comprehensive assessment of the risk of UIA instability.^22^

More small UIAs have been effectively managed in recent decades using surgical clipping and endovascular techniques. A scoring system has been recommended to improve treatment decision-making for small UIAs measuring less than 7 mm. This system considers factors such as size, multiple aneurysms, anatomical location, family history of aneurysms, age, smoking history, and aneurysm shape. A retrospective review of the dataset revealed that nearly two-thirds of the treated aneurysms had a score of more than 2, while three-fourths of the non-treated aneurysms under 7 mm had a score of 2 or lower.^36^ The aneurysm recanalization stratification scale was developed based on specific factors related to aneurysms and treatment methods. Aneurysm-specific factors included aneurysm size at the cutoff of 10 mm, rupture, and presence of thrombus. Treatment-related factors included stent assistance, flow diversion, and embolization occlusion scale. This score is more effective than Raymond Roy’s occlusion classification for predicting the need for retreatment after endovascular therapy. However, this score does not apply to small UIAs less than 10 mm and does not consider the aneurysm vessel wall information.^37^ Our study introduced a scoring prediction model for recanalization after SAC, which incorporated aneurysm-specific factors, treatment-related factors, and the recently approved imaging technique for the aneurysm wall. Although this new scale shows excellent discrimination for small UIAs less than 10 mm, it requires external validation in more extensive studies.

This study has some limitations. First, the subjects were drawn from a single center, and the sample size was relatively small, which could introduce selection bias. Additionally, the association of AWE with recanalization during long-term follow-up remains unclear, as our study had an average angiographic follow-up period of only 12 months. Furthermore, the relationship between pretreatment AWE and aneurysm recanalization resulting from increased inflammatory responses could be insufficiently explored without histological examination.

## Conclusions

Subjective assessment using CAWE and objective quantification with CRstalk on HR-VWI serve as valuable markers for aneurysm vessel wall inflammation and can predict recanalization in small UIAs following SAC. A scoring-based prediction model has been developed to evaluate the risk of recanalization, incorporating factors of aneurysm size, type of endovascular treatment, embolization occlusion grade, and CAWE. The proposed recanalization risk scale requires further investigation with a larger sample size.

## Data Availability

The original contributions presented in the study are included in the article, further inquiries can be directed to the corresponding author.

## Author contribution statement

Z-SS participated in the design of the present study. All authors participated in the interpretation and collection of the data. Q-YJ, X-LR and Z-SS wrote the initial manuscript. Z-SS revised the manuscript. All authors critically reviewed and edited the manuscript and approved the final version.

## Ethics statements

The study was reviewed and approved by the Ethics Committee of Sun Yat-sen Memorial Hospital, Sun Yat-sen University.

